# Safety profile of COVID-19 vaccines in pregnant and postpartum women in brazil

**DOI:** 10.1101/2021.12.14.21267777

**Authors:** Yaping Qiao, Ariane de Jesus Lopes de Abreu, Carolina Zampirolli Dias, Xing Meng, Rafaela Vansan Ferreira, Ramon Gonçalves Pereira, Guilherme Silva Julian, Weidong Yin

## Abstract

**Background:** Although COVID-19 vaccines are currently under use in pregnant and postpartum women, there is still lack of evidence regarding safety and effectiveness in these populations. This study aims to describe the safety profile of COVID-19 vaccines in pregnant and postpartum women in the early stage of vaccination campaign in Brazil.

**Methods:** This is an observational cross-sectional study using data from the Brazilian surveillance information system for adverse events (SI-EAPV) to characterize the safety of COVID-19 vaccines available (Sinovac/Butantan, Pfizer/BioNTech, AstraZeneca and Janssen) in Brazilian pregnant and postpartum women after receiving it from April to August 2021. A descriptive analysis was performed to assess the frequency and incidence rate of the adverse events (AE) for COVID-19 vaccines.

**Results:** A total of 3,333 adverse events following COVID-19 immunization were reported for the study population in the SIEAPV. The incidence of AE found was 309.4/100,000 doses (95% CI 297.23, 321.51). Regarding the four vaccines available in the country, Sinovac/Butantan had the lowest incidence (74.08/100,000 doses; 95% CI 63.47, 84.69). Systemic events were the most frequent notified for the group (82.07%), followed by local (11.93%) and maternal (4.74%), being most of them classified as non-severe (90.65%).

**Conclusion:** A similar pattern of AE as stated in other studies was found, with even better results for non-viral vector vaccines, corroborating to the recommendation of vaccination for these groups. Even though, further studies appraising a longer observation time are still needed to provide a broader safety aspect for the vaccines currently under use for this population.

## Introduction

The Coronavirus disease 2019 (COVID-19) has been shown to be less lethal than previous coronavirus diseases, although it is highly contagious. Also, a higher risk of severe disease has been associated with aging and comorbidities.^1^ Equally important increased risk has been noted in pregnant and postpartum women, making them particularly vulnerable to COVID-19.^1^ Studies have shown that, when compared to non-pregnant, pregnant women might develop more severe symptoms, being at increased risk of requiring hospitalization in intensive care unit, along with invasive ventilation, extra corporeal membrane oxygenation and mortality.^2,3^ In Brazil, more than 18 thousand cases of severe acute respiratory syndrome (SARS) by COVID-19 were recorded in pregnant and postpartum women, resulting in almost 1,500 deaths by June 2021.^4^ Likewise, a 20% increase in maternal mortality rate was observed in 2020.^4^

Until now, there is still lack of evidence regarding safety and efficacy of the vaccines in pregnant and postpartum women, since they were not in included in initial studies of COVID-19 vaccines.^5^ Even though, considering their higher susceptibility to COVID-19, vaccination for this group has been conducted by assessing risks and benefits.^6^ COVID-19 immunization started on January 2021 in Brazil and, in March 2021, pregnant and postpartum women with comorbidities were defined as priority group.^7^ In April 2021, Ministry of Health (MoH) recommended that this subgroup should be vaccinated, as long as a careful assessment was carried out with the physician, regardless of the gestational age.^7^ Due to adverse events experience by this subgroup, in May 2021 vaccination was changed again only for those women with comorbidity, and, in July 2021, changed to include the entire maternal population.^7^ Four COVID-19 vaccines were initially recommended - Sinovac/Butantan, Janssen, AstraZeneca and Pfizer/BioNTech, although after May 2021 there was a recommendation to remain only Sinovac/Butantan and Pfizer/BioNTech vaccines for this group^8,9^

By November 2021, about 1,7 million doses have been administered in this group, with an estimative to vaccinate more than 2,5 million pregnant and postpartum women in the country.^8,10^ Post authorization safety studies are a way to provide more evidence for this population. However, up to now, there are few evidence regarding the safety profile of those vaccines for pregnant and postpartum women from real world evidence perspective, considering pharmacovigilance systems as main source of information, especially for low and middle-income countries (LMIC), such as Brazil.^11^ In that way, this study aims to describe the incidence of adverse events (AE) reported by pregnant and postpartum women after receiving vaccines approved for use in the early stage of vaccination campaign (April 2021 to August 2021) in Brazil.

## Methods

### Surveillance Systems and Covered Population

In Brazil, records of adverse events following immunization (AEFI) in vaccinated individuals in the public services are made available by the General Coordination of the National Immunization Program (NIP). The NIP is responsible for the registration, investigation and causality analysis of AEFI reported by the public health system.^12^ For this study we requested the SI-EAPV (AEFI Surveillance Information System) dataset to the Brazilian MoH, using the Fala BR platform (https://www.gov.br/acessoainformacao/pt-br/falabr).^13^ This system is linked to the national system for reporting AE related to the use of drugs and vaccines in VigiMed, adopted at the end of 2018 by Anvisa, as a result of its partnership with the Uppsala Monitoring Centre (UMC).^14^ The SI-EAPV has the purpose to systematically monitor the notifications, investigate and consolidate data relating to AEFI occurring at the National, State, Regional, Municipal and local levels, contributing to improve the safety in the use of immunizations, with a passive surveillance approach. Following the stablished flow, during the COVID-19 pandemic, all AEFI related to COVID-19 vaccines have been notified in SI-EAPV.^15^ To assess the total number of vaccines doses administered in the country, the National Vaccination Campaign against COVID-19 database *(“Campanha Nacional de Vacinação contra a COVID-19”*), from OpenDatasus, were used.^16^ This dataset is updated daily and, for this study, we used data from 3 November 2021. Pregnant women were identified in the datasets as those who reported to be pregnant at the time they received the vaccine and postpartum were considered those women who reported to be breastfeeding in the SI-EAPV and who declared to be at postpartum at the moment of the vaccination.

Registries of AEFI with more than 50% of variables with missing data were excluded from the study. Quality check procedures within the dataset were performed; this led to the exclusion of São Paulo State from the analysis to minimize potential selection and information bias since the data was underreported.

### Study setting and outcomes

We analyzed AEFI notifications reported by pregnant and postpartum women who received any COVID-19 vaccine authorized and available in Brazil for use. AEFI could be reported as adverse event (AE) or immunization error (IE), after receiving at least one dose of a COVID-19 vaccine, including: CoronaVac (Sinovac/Butantan), Ad26.COV2.S (Janssen), ChAdOx1 nCoV-19 and BBV152 (AstraZeneca) and/ or BNT162b2 (Pfizer/BioNTech).

Demographic characteristics were described according to the age, race/ethnicity, region of the country where the notification was reported and the maternal situation of the woman. The AE were described according to the type (local, systemic and maternal)^17^, severity (severe and non-severe) as well as the case evolution (death, under investigation, cure without sequelae, unknown/ loss of follow-up and under investigation). AE reported as “COVID-19”, “PCR positive to COVID-19” and the like were classified as inconclusive and those reported as “vaccination error”, “inadvertent exposure to vaccine”, “contraindication” was classified as inconsistent.

Additionally, considering that MoH changed the recommendation to vaccinate this groups only with vaccines that do not contain viral vector, the same woman could have received different vaccines as first and second dose.^7,8^

### Statistical Analysis

Description of AE notifications characteristics were performed for women reporting to be pregnant or at postpartum after receiving a COVID-19 vaccine from April to August 2021. AE were presented as number and frequency (%) for the outcomes of interest. The incidence rate (IR) of AE per 100,000 doses applied was also estimated with 95% confidence interval (CI). IR was calculated dividing the number of AE notified during the period of the study by the number of doses administered in the same group in the same period. Data analyses were conducted using Python version 3.6.5 (Python Software Foundation).

Datasets used were public and anonymized, protecting the confidentiality and privacy of all patients.^18^ All activities were conducted according to the applicable federal laws.^19^

## Results

From April to August 2021, a total of 3,333 AEFI reported by Brazilian pregnant and postpartum women who received COVID-19 vaccines in the SI-EAPV were included in the study. Of those, 473 were from women who received Sinovac/Butantan, 788 Pfizer/BioNTech, 2,016 AstraZeneca and 56 Janssen (Figure 1). AE was the most common reported AEFI by this population (74.59%). Regarding IE, they were reported in 25.29% of the notifications, being more frequent among pregnant and postpartum women who received Sinovac/Butantan and Pfizer/BioNTech vaccines (60.47% and 73.21%, respectively) (Supplementary material 1).

**Figure 1.**
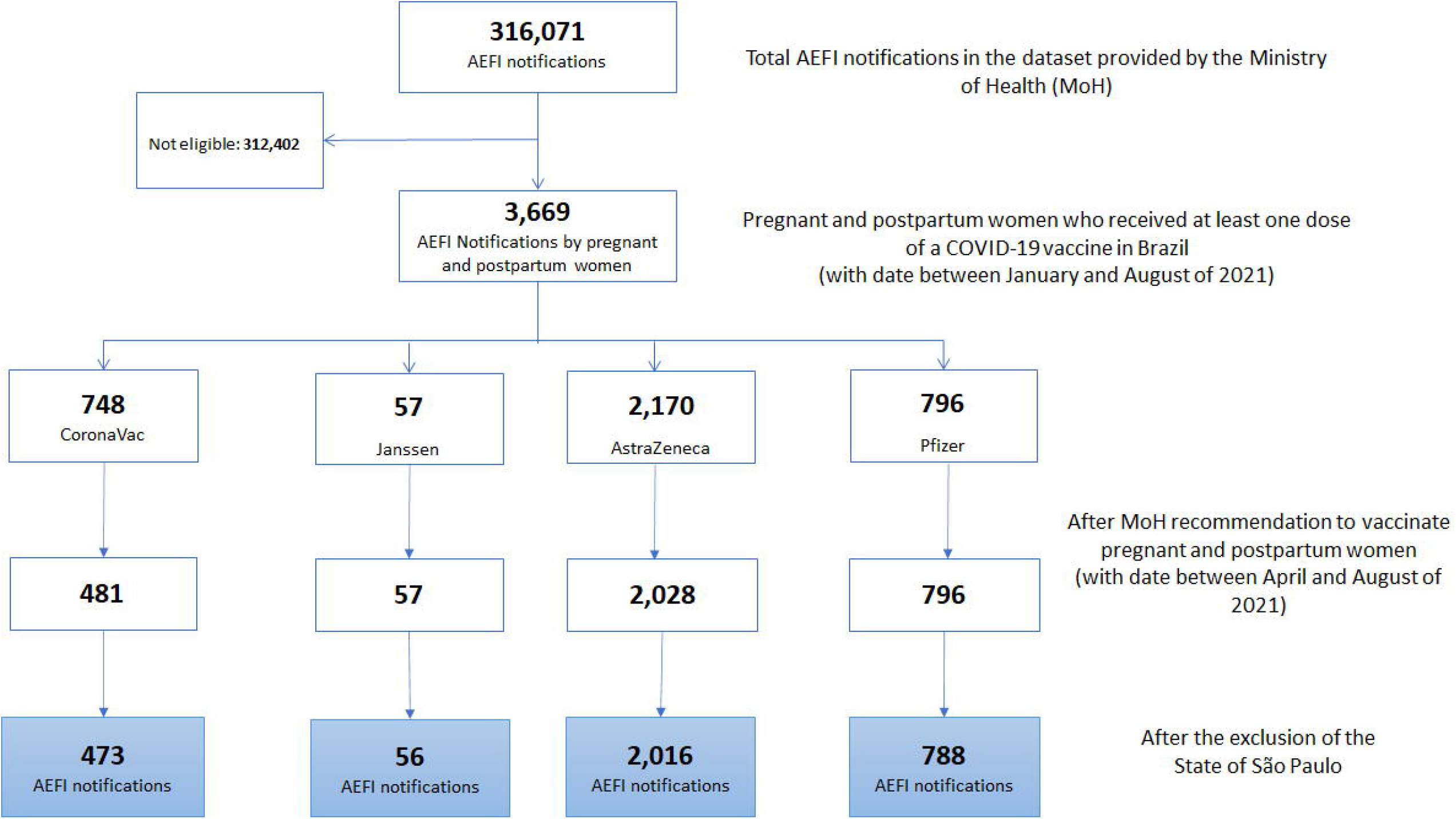
Attrition diagram

### Adverse Events reported in SIEAPV

AE notifications were more frequent among pregnant and postpartum women aged 20 to 35 years old, with a mean age of 28.56 (Standard Deviation 7.2), who reported as white (42.84%) and brown (36.89%) (Table 1). From a regional perspective, even excluding the state of São Paulo, the South (34.31%) and Southeast (33.79%) regions had most part of the notification from pregnant and postpartum women, although it changed according to the vaccine received: for Sinovac/Butantan the highest were noted in the South (40.64%), for Pfizer/BioNTech in the Southeast (45.45%), for AstraZeneca in the South (35.75%) and for Janssen in the Northeast (46.67%). According to the maternal situation, few women reported as being at postpartum (1.53%) and the distribution within the three trimesters of pregnancy were similar (25.34% in the first, 30.01% in the second and 32.82% in the third trimester of pregnancy) (Table 1).

**Table 1.**
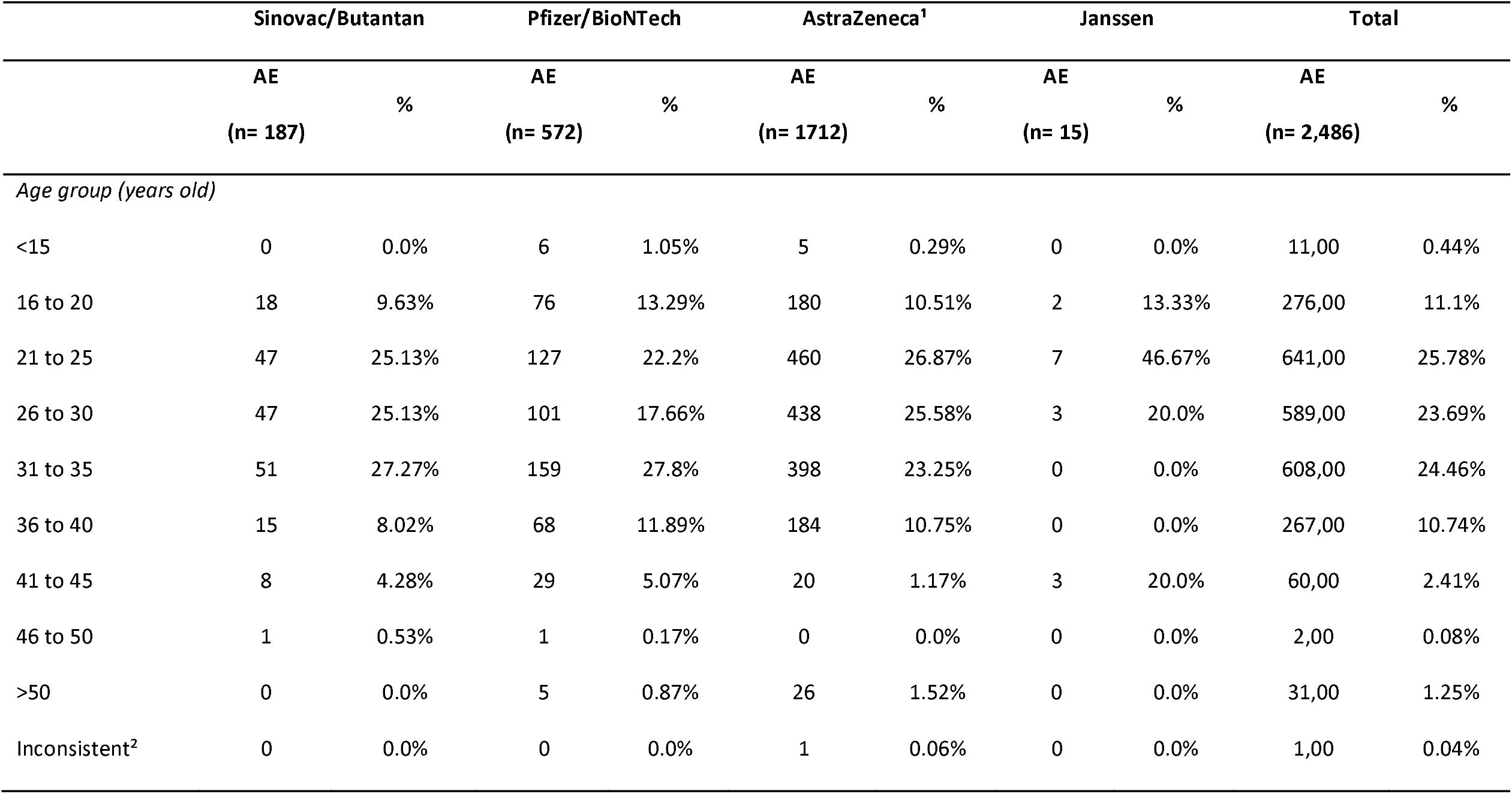
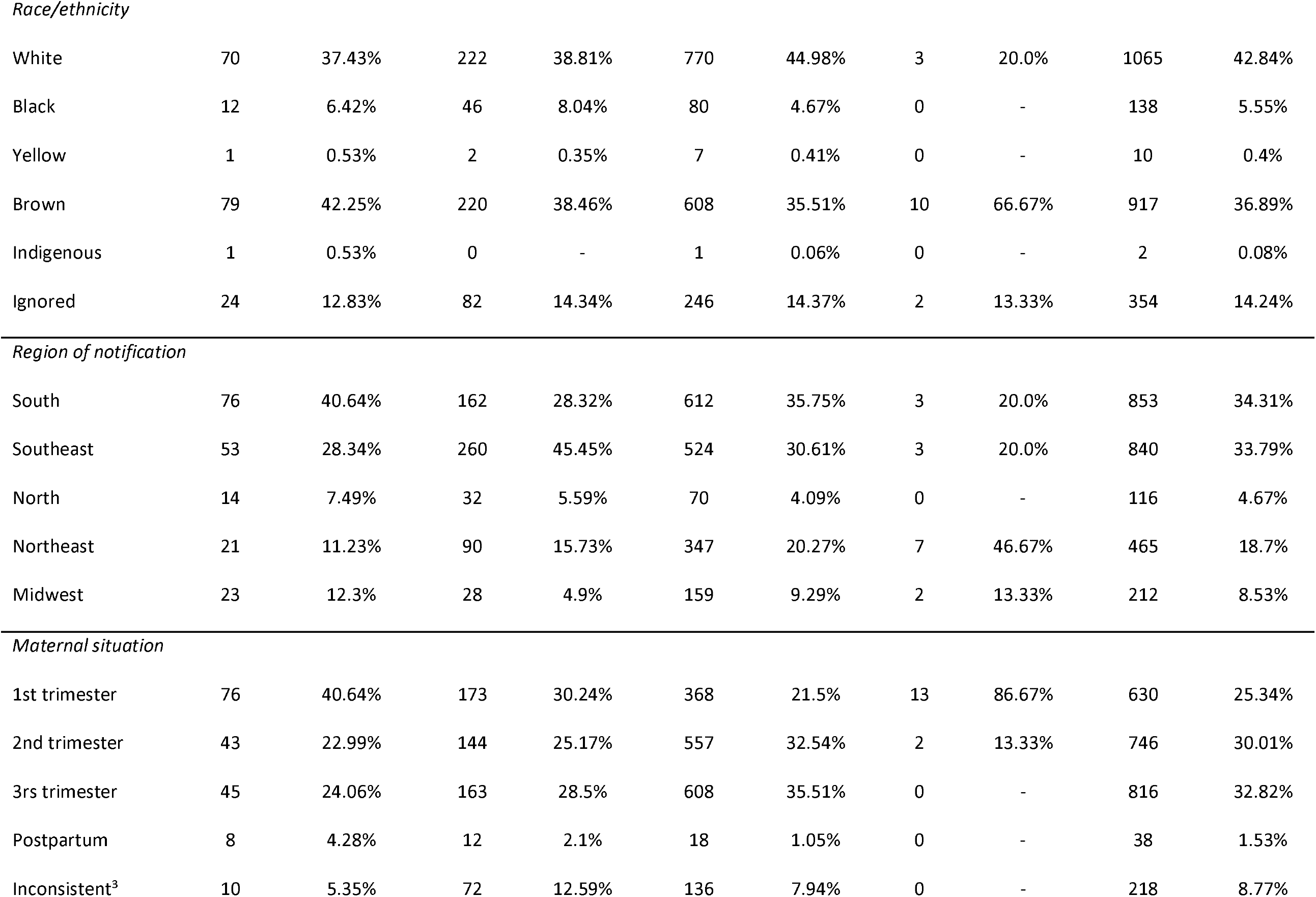
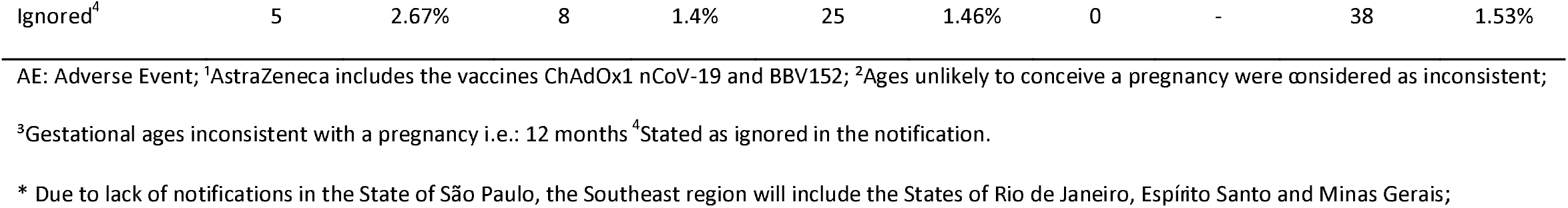
Sociodemographic characteristics of pregnant and postpartum women that notified adverse events after receiving vaccines against COVID-19

### Incidence of adverse events

The overall incidence of AE among pregnant and postpartum women was 309.4 / 100,000 doses of vaccines administered (95% CI 297.23, 321.51). In the analysis according to the maternal situation, IR by pregnant women was 404.3 / 100,000 doses (95% CI 388.75, 420.75) and by postpartum 19.6 / 100,000 doses (95% CI 13.47, 25.78). Regarding the four vaccines available in the country, Sinovac/Butantan vaccine had the lowest IR (74.08 / 100,000 doses; 95% CI 63.47, 84.69) (Table 2).

**Table 2.** Incidence of adverse events notified by Brazilian pregnant and postpartum women by vaccine received

|  | Overall |  |  |  | Pregnant |  |  |  | Postpartum |  |  |  |
| --- | --- | --- | --- | --- | --- | --- | --- | --- | --- | --- | --- | --- |
|  | AE | N<br>doses | IR* | 95% CI | AE | N<br>doses | IR* | 95% CI | AE | N<br>doses | IR* | 95% CI |
| Sinovac/Butantan | 187 | 252430 | 74.08 | (63.47, 84.69) | 179 | 193517 | 92.5 | (78.95, 106.04) | 9 | 58913 | 15.28 | (5.3, 25.26) |
| Pfizer/BioNTech | 572 | 484310 | 118.11 | (108.43, 127.78) | 560 | 372980 | 150.14 | (137.72, 162.57) | 12 | 111330 | 10.78 | (4.68, 16.88) |
| AstraZeneca<sup>1</sup> | 1712 | 65435 | 2616.34 | (2494.04, 2738.64) | 1694 | 37954 | 4463.3 | (4255.55, 4671.04) | 18 | 27481 | 65.5 | (35.25, 95.75) |
| Janssen | 15 | 1388 | 1080.69 | (536.76, 1624.62) | 15 | 367 | 4087.19 | (2061.54, 6112.85) | 0 | 1021 | 0 | (0.0, 0.0) |
| Total | 2486 | 803563 | 309.37 | (297.23, 321.51) | 2448 | 604818 | 404.75 | (388.75, 420.75) | 39 | 198745 | 19.62 | (13.47, 25.78) |

AE: Adverse Events; IR: incidence rate; CI: Confidence interval;
\* IR per 100,00 doses; <sup>1</sup>AstraZeneca includes the vaccines ChAdOx1 nCoV-19 and BBV152;
N.B: Pregnant women can be breastfeeding; hence, they will count in pregnant and postpartum columns.

Stratifying the AE according to type, among pregnant women, systemic events were the most frequent notified (82.03%), followed by local (11.93%) and maternal (4.78%), being most of them classified as non-severe (90.65%) (Supplementary appendix 2). Also, the IR of systemic events in the Brazilian pregnant women was the highest (249.88/ 100,000 doses: 95% CI 238.97, 260.8). Maternal AE IR was 14.56/ 100,000 doses (95% CI 11.92, 17.2) (Table 3).

**Table 3.** Incidence of AE among pregnant women according to the type and severity

|  | Sinovac/Butantan |  | Pfizer/BioNTech |  | AstraZeneca<sup>1</sup> |  | Janssen |  | Total |  |
| --- | --- | --- | --- | --- | --- | --- | --- | --- | --- | --- |
|  | IR* | 95% CI | IR* | 95% CI | IR* | 95% CI | IR* | 95% CI | IR* | 95% CI |
| Maternal | 10.3 | (6.34, 14.26) | 9.7 | (6.93, 12.48) | 65.71 | (46.08, 85.35) | 72.05 | (0.0, 213.2) | 14.56 | (11.92, 17.2) |
| Systemic | 50.31 | (41.56, 59.06) | 89.41 | (80.99, 97.82) | 2194.5 | (2082.29, 2306.8) | 864.55 | (377.51, 1351.59) | 249.88 | (238.97, 260.8) |
| Local | 4.36 | (1.78, 6.93) | 15.49 | (11.98, 18.99) | 311.76 | (269.05, 354.47) | 144.09 | (0.0, 343.65) | 36.34 | (32.17, 40.51) |

IR: incidence rate; CI: confidence interval; <sup>1</sup>AstraZeneca includes the vaccines ChAdOx1 nCoV-19 and BBV152;
\*IR *per* 100,000 doses administered.

The most common maternal AE notified by pregnant and postpartum women included spontaneous abortion (2.37%) pregnancy bleeding (0.76%) and neonatal death (0.52%). Among the non-maternal AE, headache (18.54%), fever (13.79%), myalgia (10.30%) and pain (7.60%) were the most reported. The most frequent AE were similar among the four vaccines, except for pain, which was less frequent reported for those who received Sinovac/Butantan vaccine (Table 4).

**Table 4.** Frequency of most common adverse events experienced by pregnant and postpartum women receiving COVID-19 vaccines and who reported an AEFI

|  | Sinovac/Butantan |  | Pfizer/BioNTech |  | AstraZeneca<sup>1</sup> |  | Janssen |  | Total |  |
| --- | --- | --- | --- | --- | --- | --- | --- | --- | --- | --- |
|  | AE | % | AE | % | AE | % | AE | % | AE | % |
|  | (n= 187) |  | (n= 572) |  | (n= 1712) |  | (n= 15) |  | (n=2,486) |  |
| **Maternal** |  |  |  |  |  |  |  |  |  |  |
| Spontaneous abortion | 9 | 4.81% | 28 | 4.9% | 21 | 1.23% | 1 | 0.066% | 59 | 2.37% |
| Pregnancy bleeding | 6 | 3.21% | 3 | 0.52% | 11 | 0.64% | 0 | 0% | 20 | 0.80% |
| Neonatal death | 3 | 1.6% | 9 | 1.57% | 1 | 0.06% | 0 | 0% | 13 | 0.52% |
| Premature birth | 1 | 0.53% | 3 | 0.52% | 3 | 0.18% | 0 | 0% | 7 | 0.28% |
| Abdominal pregnancy | 3 | 1.6% | 0 | 0% | 0 | 0% | 0 | 0% | 3 | 0.12% |
| ***Non-maternal*** |  |  |  |  |  |  |  |  |  |  |
| Headache | 20 | 10.7% | 77 | 13.46% | 360 | 21.03% | 4 | 28.57% | 461 | 18.54% |
| Fever | 13 | 6.95% | 46 | 8.04% | 282 | 16.47% | 2 | 14.29% | 343 | 13.8% |
| Myalgia | 12 | 6.42% | 37 | 6.47% | 206 | 12.03% | 1 | 7.14% | 256 | 10.3% |
| Pain | 3 | 1.6% | 35 | 6.12% | 149 | 8.7% | 2 | 14.29% | 189 | 7.6% |
| Vomit | 2 | 1.07% | 20 | 3.5% | 67 | 3.91% | 2 | 14.29% | 91 | 3.66% |
| Chills | 4 | 2.14% | 8 | 1.4% | 77 | 4.5% | 0 | 0% | 89 | 3.58% |
| Nausea | 2 | 1.07% | 19 | 3.32% | 62 | 3.62% | 1 | 7.14% | 84 | 3.38% |
| Abdominal pain | 9 | 4.81% | 22 | 3.85% | 44 | 2.57% | 1 | 7.14% | 76 | 3.06% |
| Diarrhea | 3 | 1.6% | 20 | 3.5% | 40 | 2.34% | 0 | 0% | 63 | 2.53% |
| Arm Pain | 0 | 0% | 26 | 4.55% | 34 | 1.99% | 0 | 0% | 60 | 2.41% |
| Asthenia | 6 | 3.21% | 15 | 2.62% | 35 | 2.04% | 0 | 0% | 56 | 2.25% |
| Fatigue | 2 | 1.07% | 14 | 2.45% | 39 | 2.28% | 0 | 0% | 55 | 2.21% |
| Cough | 7 | 3.74% | 28 | 4.9% | 17 | 0.99% | 0 | 0% | 52 | 2.09% |
| Dyspnea | 5 | 2.67% | 15 | 2.62% | 31 | 1.81% | 0 | 0% | 51 | 2.05% |
| Runny nose | 7 | 3.74% | 23 | 4.02% | 16 | 0.93% | 0 | 0% | 46 | 1.85% |
| Sore throat | 4 | 2.14% | 11 | 1.92% | 22 | 1.29% | 0 | 0% | 37 | 1.49% |
| Edema | 4 | 2.14% | 9 | 1.57% | 14 | 0.82% | 0 | 0% | 27 | 1.09% |

AE: Adverse event; <sup>1</sup>AstraZeneca includes the vaccines ChAdOx1 nCoV-19 and BBV152.

Regarding the case evolution of the events, 53.30% were missing and 30.85% reported as being cured without sequelae (Supplementary material 3).

## Discussion

In the COVID-19 pandemic reality, despite the recognition of the need for inclusion of pregnant and postpartum women in clinical trials, the speed at which the COVID-19 vaccines were developed, and trials conducted precluded inclusion of them.^20^ In that sense, post authorization safety studies are a way to provide more evidence for this population.

Using surveillance data, we found more than 3,000 events notifications by pregnant and postpartum women after receiving at least one dose of a COVID-19 vaccine in the early stage of the campaign in Brazil. AE were the most common, although for some vaccines IE were more frequent – which may reflect the changes in recommendation to vaccinate this group disposed by the Brazilian MoH.^8,21^

Concerning the frequency of AE, the distribution according to the age and race/ethnicity was similar within all vaccines available. However, in a regional perspective, it differed as each region/state could have differences in the cold chain distribution strategies.^8^ Among the maternal population, pregnant women were responsible for most of the notifications reported in the period.

The overall incidence of AE found for this population was 309.4/ 100,000 doses. Although there is a lack of evidence regarding the safety of these vaccines in the maternal population, our findings are in accordance with the available literature for other studies that assessed safety of COVID-19 in different populations groups. An epidemiological bulletin from the Brazilian MoH from January to February of 2021 showed a 350.9 AEFI notifications/ 100,000 doses of vaccines early administered in the Brazilian population.^22^ Another study conducted in Minas Gerais state that assessed safety of COVID-19 vaccines from January to March 2021 found an incidence rate of 777.12 AEFI per 100,000 doses applied, with 97% of them classified as non-severe AE.^23^

In relation to the magnitude of our findings when compared to other vaccines recommended to pregnant and postpartum women in Brazil, a study from Silveira IO *et al*, assessing adverse events from the SI-EAPV database from 2015 to 2019 in Minas Gerais state, found an overall incidence of 76.9 AEFI notifications/100,000 doses.^24^ Findings related to race/ ethnicity, type of event and case evolution were also similar to the patterns found in our results.^24^

As for the systemic events found in our study, the most frequent types follow a similar pattern described by Gattás VL, et al in relation to the ones found for influenza vaccine, which is recommended for any gestational age in Brazil, as COVID-19 vaccines are.^25^ Although we have not compared pregnant to non-pregnant women, there are studies suggesting that the physiologic changes in pregnancy seems to not materially affect non-maternal events.^26,27^

Additionally, our study describes with more emphasis systemic events classified as maternal, showing an incidence of 14.56 AE notifications/ 100,000 doses, of which spontaneous abortion was the most frequent type of event (2.37%) and with differences in the frequency found for the different types of vaccines available. Brazilian data up to 2019 showed a proportion of 3.4% of spontaneous abortion in the country. Meanwhile, spontaneous abortion incidence in the literature varies from 6.5% to 21% of pregnancies, and it is recognized as one of the most common complications during a pregnancy.^28–30^ In addition, a study assessing safety of mRNA COVID-19 vaccines in pregnant population in the United States assessed by the V-Safe pregnancy registry system found an overall frequency of spontaneous abortion of 12.6% among pregnant women who received a COVID-19 vaccines.^26^ Cardoso BB et al^31^, however, argues that data on abortion and its complications may be incomplete in Brazil, since the hospitalizations occurred due to an abortion is only one data source to estimate the total number of abortions in the country.^31^

When comparing the different vaccines administered in this population in Brazil, we found that Sinovac/Butantan and Pfizer/BioNTech vaccines had the lowest IR of AE, which is in line with MoH recommendations to only administer them in pregnant and postpartum woman.^8,32^

Our study has some limitations. The AEFI notifications used in this study are subject to limitations of passive surveillance system. This means that each health level routinely and periodically sends information about the events subject to surveillance at the immediately superior. In the same way, the classifications in the database might be susceptible to the interpretation of the person filling out the system, implying in the possibility of lack of uniformity in reporting the characteristics of the event and in the place where the information is filled in the form of the surveillance system. Although these systems generate valuable information regarding the description of the occurrence of adverse events, they usually do not allow establishing causality between the occurrence of AEFI and the vaccine.^11,33–35^ In that sense, our study is unable to evaluate AE outcomes that might occur in association with exposures earlier in pregnancy or postpartum period.

Furthermore, the definition of postpartum women varied in the data sources used - which may underestimate the incidence in this population. In the same direction, the Brazilian obstetric observatory for COVID-19 has been showing that there are inconsistencies in the vaccination information fulfilling, since they found pregnant and postpartum women of male sex, and AEFI notified for COVID-19 vaccines administered previous to the vaccination campaign start and over 55 years old.^4^

Another potential limitation of our analyses is the underreporting of AE. SIEAPV is a passive surveillance system. Underreporting can occur due to difficulties in the conclusion of cases investigations and in adherence of the population to notify the events. In that sense, mild to moderate events might be more underreported than severe that required hospitalization or more intensive care. On the other hand, each of the four vaccines were available in different moments (Sinovac/Butantan and AstraZeneca since January, Pfizer/BioNTech since May and Janssen since July 2021).^8^ The Janssen vaccine was first provided to the population when there was already the recommendation not to vaccinate pregnant and postpartum women with viral vector vaccines, which led to a small number of doses administered in this population, hence, a low number of notifications up to the cut-off period of this study. In the same sense, Sinovac/Butantan and AstraZeneca vaccines were available in the beginning of the campaign, when only women with comorbidities were being vaccinated.^8^ Also, some of the AE presented in this study might be still under investigation during the study period and might not have a final classification.

Nevertheless, our study allows a better understand of COVID-19 vaccines safety profile under a vaccination campaign placed during a pandemic setting in a LMIC as Brazil. We found a similar pattern of AE as stated in other studies, with even better results for non-viral vector vaccines, corroborating that vaccination of this groups should continue as a priority. Further studies appraising a longer time for a better understanding adverse events incidence in relation to second and booster doses and the component of vaccine interchangeability are still needed to provide a broader safety aspect for the vaccines currently under use for this population.

## Data Availability

All data produced are available online at Fala BR platform and Opendatasus websites.

https://www.gov.br/acessoainformacao/pt-br/falabr

https://opendatasus.saude.gov.br/

## Contributors

AJLA, YPQ and GSJ conceptualized the study. RGP, RVF, AJLA and CZD analyzed the data. RGP and RVF curated the data. AJLA, YPQ, CZD and GSJ wrote the first draft of the manuscript. YPQ, XM, WDY and other authors reviewed and edited revisions of the manuscript, had full access to all the data in the study, and had final responsibility for the decision to submit for publication.

