## Supplemental tables 2 and 3 and supplemental figure 1 for "Safety profile of COVID-19 vaccines in pregnant and postpartum women in brazil"

### Supplementary material

S1. Frequency of adverse events and immunization error among all AEFI notified

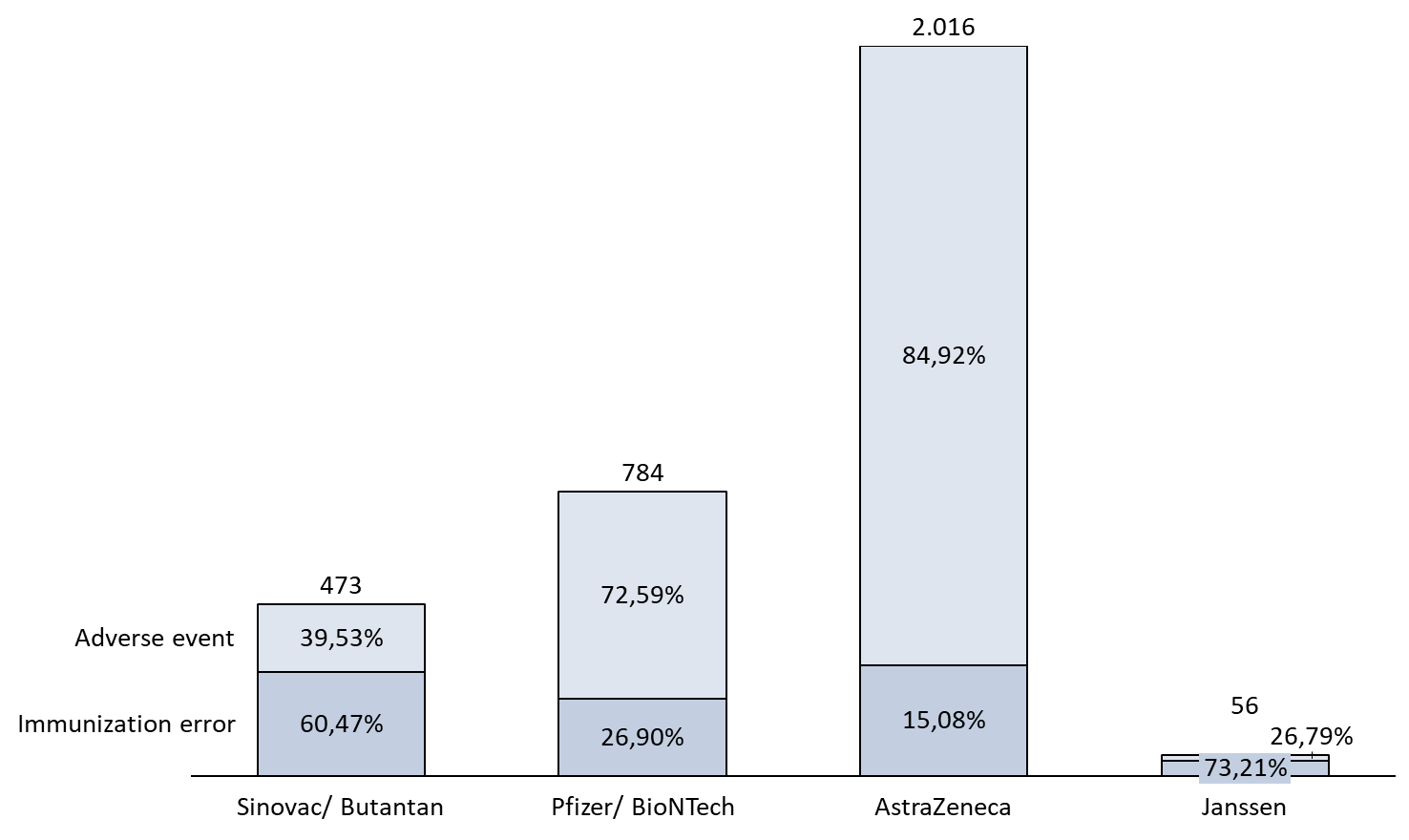

*AstraZeneca includes the vaccines ChAdOx1 nCoV-19 and BBV152.

S2. Frequency of adverse events according to the type and severity

|  | **Pregnant** | | | | | | | | | | **Postpartum** | | | | | | | | | |
| --- | --- | --- | --- | --- | --- | --- | --- | --- | --- | --- | --- | --- | --- | --- | --- | --- | --- | --- | --- | --- |
|  | **Sinovac/ Butantan** | | **Pfizer/ BioNTech** | | **AstraZeneca¹** | | **Janssen** | | **Total** | | **Sinovac/ Butantan** | | **Pfizer/ BioNTech** | | **AstraZeneca¹** | | **Janssen** | | **Total** | |
|  | **AE** | **%** | **AE** | **%** | **AE** | **%** | **AE** | **%** | **AE** | **%** | **AE** | **%** | **AE** | **%** | **AE** | **%** | **AE** | **%** | **AE** | **%** |
| *Adverse Event Type* | | | | | | | | | | | | | | | | | | | | |
| Maternal | 26 | 14.53% | 47 | 8.39% | 43 | 2.53% | 1 | 6.67% | 117 | 4.78% | 0 | 0 | 0 | 0 | 0 | 0 | - | - | 0 | 0 |
| Local | 11 | 6.15% | 75 | 13.39% | 204 | 12.04% | 2 | 13.33% | 292 | 11.93% | 0 | 0 | 2 | 16.67% | 1 | 5.56% | - | - | 3 | 7.69% |
| Systemic | 127 | 70.95% | 433 | 77.32% | 1436 | 84.76% | 12 | 80.0% | 2008 | 82.03% | 7 | 77.78% | 10 | 83.33% | 17 | 94.44% | - | - | 34 | 87.18% |
| Inconclusive* | 13 | 7.26% | 5 | 0.89% | 9 | 0.53% | 0 | 0 | 27 | 1.1% | 2 | 22.22% | 0 | 0 | 0 | 0 | - | - | 2 | 5.13% |
| Inconsistent** | 2 | 1.12% | 0 | 0 | 2 | 0.12% | 0 | 0 | 4 | 0.16% | 0 | 0 | 0 | 0 | 0 | 0 | - | - | 0 | 0 |
| Severe | 36 | 20.11% | 96 | 17.14% | 93 | 5.49% | 1 | 6.67% | 226 | 9.23% | 4 | 44.44% | 1 | 8.33% | 0 | 0 | - | - | 5 | 12.82% |
| Non-severe | 143 | 79.89% | 463 | 82.68% | 1599 | 94.39% | 14 | 93.33% | 2219 | 90.65% | 5 | 55.56% | 11 | 91.67% | 18 | 100.0% | - | - | 34 | 87.18% |
| Ignored | 0 | 0 | 1 | 0.18% | 2 | 0.12% | 0 | 0 | 3 | 0.12% | 0 | 0 | 0 | 0 | 0 | 0 | - | - | 0 | 0 |

AE: Adverse Events; ¹ AstraZeneca includes the vaccines ChAdOx1 nCoV-19 and BBV152;

*AE reported as “COVID-19”, “PCR positive to COVID-19” and the like were classified as inconclusive;

**AE reported as “vaccination error”, “inadvertent exposure to vaccine”, “contraindication” was classified as inconsistent.

S3. Case evolution of the adverse events notifications according to the vaccine types

|  | **Sinovac/ Butantan** | | **Pfizer/ BioNTech** | | **AstraZeneca¹** | | **Janssen** | | **Total** | |
| --- | --- | --- | --- | --- | --- | --- | --- | --- | --- | --- |
|  | **AE (n= 187)** | **%** | **AE (n= 572)** | **%** | **AE (n= 1712)** | **%** | **AE (n= 15)** | **%** | **AE (n= 2486)** | **%** |
| Death | 8 | 4.28% | 21 | 3.67% | 10 | 0.58% | 0 | 0 | 39 | 1.57% |
| Under investigation | 33 | 17.65% | 95 | 16.61% | 205 | 11.97% | 1 | 6.67% | 334 | 13.44% |
| Cure without sequelae | 36 | 19.25% | 97 | 16.96% | 628 | 36.68% | 6 | 40.0% | 767 | 30.85% |
| Cure with sequelae | 0 | 0 | 8 | 1.4% | 12 | 0.7% | 1 | 6.67% | 21 | 0.84% |
| Missing | 110 | 58.82% | 351 | 61.36% | 857 | 50.06% | 7 | 46.67% | 1325 | 53.3% |

¹AstraZeneca includes the vaccines ChAdOx1 nCoV-19 and BBV152.
